# Pharmacogenomics of hypertension in chronic kidney disease: the CKD-PGX study

**DOI:** 10.1101/2021.03.30.21254665

**Authors:** Michael T. Eadon, Judith Maddatu, Sharon M. Moe, Arjun D. Sinha, Ricardo Melo Ferreira, Brent W. Miller, S. Jawad Sher, Jing Su, Victoria M. Pratt, Arlene B. Chapman, Todd S. Skaar, Ranjani N. Moorthi, for the CKD-PGX investigators

## Abstract

**Background:** Patients with chronic kidney disease (CKD) often have uncontrolled hypertension despite polypharmacy. Pharmacogenomic drug-gene interactions (DGIs) may impact the metabolism or efficacy of antihypertensive agents. We hypothesized that providing a panel of 11 pharmacogenomic predictors of antihypertensive response would improve hypertension control.

**Methods:** A prospective cohort with CKD and hypertension was followed to assess the effect of pharmacogenomic testing on blood pressure control. The analysis population included 382 hypertensive subjects genotyped for cross-sectional assessment of drug-gene interactions and 335 subjects followed for 1 year to assess systolic (SBP) and diastolic blood pressure (DBP).

**Results:** Most participants (58.2%) with uncontrolled hypertension had a DGI reducing the efficacy of one or more antihypertensive agents. Subjects with a DGI had 1.88-fold (95% CI 1.2-2.8) higher odds of uncontrolled hypertension as compared to those without a DGI, adjusted for race and CKD grade. *CYP2C9* reduced metabolism genotypes were associated with losartan response and uncontrolled hypertension (Odds Ratio 5.2, CI 1.9 -14.7). *CYP2D6* intermediate or poor metabolizers had less frequent uncontrolled hypertension compared to normal metabolizers taking metoprolol or carvedilol (OR 0.55, CI 0.3-0.95). In 335 subjects completing 1 year follow-up, SBP (−4.0 mmHg, CI 1.6-6.5) and DBP (−3.3 mmHg, CI 2.0-4.6) were improved. The magnitude of reductions in SBP (−14.8 mmHg, CI 10.3-19.3) and DBP (−8.4 mmHg, CI 5.9-10.9) were greatest in the 90 individuals with uncontrolled hypertension and an actionable genotype.

**Conclusions:** There is a potential role for the addition of pharmacogenomic testing to optimize antihypertensive regimens in patients with CKD.

## INTRODUCTION

Hypertension and chronic kidney disease (CKD) are common intersecting diseases with enormous economic burden, morbidity, and mortality. The Center for Disease Control (CDC) reports that 45% of the US population has hypertension, with approximately half of those individuals inadequately controlled^1^. The prevalence of hypertension increases with the severity of CKD as 36% of grade 1, 48% of grade 2, 60% of grade 3, and 84% of grade 4/5 CKD patients have concomitant hypertension^2^. Impaired sodium excretion, extracellular volume expansion, activation of the renin-angiotensin system, and numerous vasoconstrictive effects all conspire to impair blood pressure control in CKD patients^3^. International guidelines emphasize control of hypertension to reduce cardiovascular events in patients with CKD^4^.

Many antihypertensive agents are subject to different forms of pharmacokinetic or pharmacodynamic drug-gene interactions, each of which impact efficacy. For example, metoprolol is metabolized by the enzyme cytochrome P450 2D6 (CYP2D6), wherein poor metabolizers possess higher circulating concentrations of the drug at a given dose^5^. The beta-blocker class may be pharmacodynamically affected by beta-1 adrenergic receptor (ADRB1) polymorphisms^6^. The angiotensin receptor blocker (ARB) losartan potassium is a pro-drug metabolized by cytochrome P450 2C9 (CYP2C9)^7^. Poor metabolizers of CYP2C9 have lower concentrations of its active metabolite^8^. Hydralazine hydrochloride undergoes phase 2 metabolism by N-acetyltransferase 2 (NAT2)^9^. Fast and intermediate acetylators will have lower concentrations and reduced efficacy of hydralazine at a given dose. The evidence supporting these (and other) pharmacogenomic drug-gene pairs has been generated through prior clinical studies and this evidence has been previously summarized^10, 11^.

Previous studies have identified individual drug-gene pairs relevant to antihypertensives in the general population and those with CKD. Over 80% of individuals with CKD and hypertension take 2 or more antihypertensives and 32% take four or more agents^12^. Despite polypharmacy, 10.3 million Americans with apparent treatment resistant hypertension remain uncontrolled^13^.

We hypothesized that embedding a panel of pharmacogenomic predictors of antihypertensive response in routine clinical practice would aid patients and practitioners in arriving at an efficacious blood pressure regimen, either by identifying less efficacious medications in an individual’s current regimen or selecting an efficacious drug as the “next” antihypertensive agent. To facilitate this, a clinical genotyping assay was developed and implemented across multiple health systems, with results and recommendations recorded in the electronic health records (EHR) for 40 variants and 11 drug-gene pairs relevant to hypertension control^14^.

We present the results of a prospective cohort study entitled CKD-PGX, that enrolled and genotyped 382 adult hypertensive CKD patients from Indiana University Health Physicians nephrology clinics in three settings: a university health system, a county safety-net health system, and outlying suburban clinics near Indianapolis. The goal was to assess provider utilization, patient attitude, prevalence of actionable drug-gene interactions (DGIs), and blood pressure control after pharmacogenomic panel testing.

## MATERIALS AND METHODS

### Study Design

This was a prospective, observational cohort study. Subjects were recruited and provided informed consent during a nephrology clinic visit in the IU Health or Eskenazi Health systems between 2017 and 2019. Blood pressure (BP) was assessed upon enrollment and at 1 year follow-up. This study was approved by the Institutional Review Board of Indiana University (IRB # 1705413046).

### Study Population

Subjects were eligible for inclusion if of age ≥ 18 years old with the ability to provide consent and a genotyping sample. Subjects were required to have at least one of the following: systolic BP ≥ 140 mm Hg on any two readings in the 12 preceding months, estimated glomerular filtration rate (eGFR) less than 60 mL/min/1.73 m^2^, daily proteinuria > 0.2 g by 24-hour urine collection or > 0.2 g/g urine protein to creatinine ratio^15^. Our analysis population was comprised of N = 425 for the baseline subject survey, N = 382 for the cross-sectional analysis between genotype and hypertension control, and N = 335 for the longitudinal 1-year follow-up blood pressure outcomes. The prevalence of hypertension in those with CKD presenting to our clinics during the study period determined the sample size.

### Study Procedures

BP was obtained at baseline immediately following a nephrology clinic appointment using a standard sphygmomanometer while seated at rest and again from the clinic visit closest to 1 year after enrollment (± 6 months). Three BP measurements were acquired, each separated by 5 minutes. Systolic and diastolic BP were each averaged separately for the three measurements. Participants provided a whole blood or saliva sample and were genotyped for 40 variants in 11 genes related to antihypertensive response (Table 1, Supplemental Table S1). Genotyping was performed on a custom Taqman™ OpenArray™ (FisherScientific, Waltham, MA) as previously described^14^. Genetic data were deposited in the EHR approximately two weeks after testing along with interpretations on drug efficacy. Providers received encrypted email alerts when the genetic information was available in the EHR.

**Table 1:**

| Gene<sup>A</sup> | Biological/Functional Significance | Genotype or metabolizer status | Predicted Phenotype | Reference(s) |
| --- | --- | --- | --- | --- |
| *CYP2C9* | Encodes cytochrome P450 2C9 which metabolizes losartan into active metabolites<sup>B</sup>. | *1/*1, *1/*2 | Standard exposure | 7, 8, 26-34 |
|  |  | *2/*2, *1/*3, *2/*3, *3/*3, *1/*8, *1/*11, *3/*8 | Reduced active metabolite exposure |  |
| *CYP2D6* | Encodes cytochrome P450 2D6, which metabolizes / inactivates metoprolol and carvedilol. | Ultrarapid Metabolizer | Reduced drug exposure | 5, 22, 35 |
|  |  | Normal Metabolizer | Standard efficacy |  |
|  |  | Intermediate or Poor Metabolizer | Increased drug exposure |  |
| *NAT2* | Encodes N-acetyltransferase-2, which acetylates hydralazine to its inactive metabolite. | Fast or intermediate acetylator | Reduced hydralazine exposure | 9 |
|  |  | Slow acetylator | Increased hydralazine exposure |  |
| *F7*<br>(rs6046)<sup>C</sup> | Encodes clotting factor VII, blood pressure effect may be related to its role in endothelial homeostasis. | G/G | Standard amlodipine efficacy | 36 |
|  |  | G/A or A/A | Reduced amlodipine efficacy |  |
| *ADRB1* | Encodes β1adrenoceptor. | 0 copies of 49S-389R | Reduced beta blocker response | 6, 37-40 |
|  |  | 1 copy of 49S-389R | Standard beta blocker response |  |
|  |  | 2 copies of 49S-389R | Greater beta blocker response |  |
| *GRK4* | Encodes G-protein coupled receptor kinase 4, which maintains ADRB1 cell surface localization. | 0 copies of 65L-142V | Greater beta blocker response | 41, 42 |
|  |  | 1 copy of 65L-142V | Standard beta blocker response |  |
|  |  | 2 copies of 65L-142V | Reduced beta blocker response |  |
| *NEDD4L*<br>(rs4149601)<sup>D</sup> | Encodes an E3 ubiquitin ligase, which regulates ENaC expression. | G/G | Increased diuretic efficacy | 43, 44 |
|  |  | G/A | Standard diuretic efficacy |  |
|  |  | A/A | Reduced diuretic efficacy |  |
| *NPHS1*<br>(rs3814995) | Encodes the principal structural protein of the glomerular podocytes, nephrin. | G/G | Standard ARB efficacy | 45, 46 |
|  |  | G/A or A/A | Increased ARB efficacy |  |
| *VASP*<br>(rs10995)<sup>D</sup> | Encodes vasodilator stimulated phosphoprotein, regulates smooth muscle contraction. | A/A | Standard thiazide efficacy | 47 |
|  |  | A/G or G/G | Increased thiazide efficacy |  |
| *YEATS4*<br>(rs7297610)<sup>C</sup> | Expression quantitative trait locus associated with thiazide efficacy in African Americans. | C/C | Standard thiazide efficacy | 23, 48 |
|  |  | C/T or T/T | Reduced thiazide efficacy |  |
| *EBF1/FGF5/SH2B3*<sup>D</sup> | Three gene model identified by GWAS but no evidence for direct functional/biological significance | 0 efficacy alleles | Reduced thiazide efficacy | 24 |
|  |  | 1 or 2 efficacy alleles | Standard thiazide efficacy |  |
|  |  | 3 or more efficacy alleles | Increased thiazide efficacy |  |

<sup>A</sup>Only antihypertensive pharmacogenomic genes are included here. For multiple single nucleotide variant models, please see the supplement for the variants tested. Additional pharmacogenomic variants unrelated to hypertension, but for genetic risk prediction genotype data was given to providers. <sup>B</sup>Not all Angiotensin receptor blockers are metabolized by CYP2C9, e.g. Olmesartan does undergo significant metabolism. <sup>C</sup>This variant only has supporting data in individuals of African ancestry. Clinicians were advised not to extrapolate findings to other populations. <sup>D</sup>This variant only has supporting evidence in individuals of European ancestry. Clinicians were advised not to extrapolate findings to other populations. ENaC – epithelial sodium channel of the principal cell, ARB – Angiotensin receptor blocker, GWAS – genome wide association study. 49S refers to the A allele of rs1801252 and 389R refers to the C allele of rs1801253.

Nephrology providers (N = 39) gave assent to enroll their patients and were trained on the interpretation of pharmacogenomic drug-gene interactions. The principal investigators did not alter or suggest changes to subjects’ prescriptions; all clinical care was at the behest of the primary nephrology provider.

Three surveys were administered: 1) each subject’s attitude toward genetic testing was evaluated in a 15 question survey at baseline (Supplemental Document S1), 2) each provider (N = 76) completed a baseline survey regarding their attitude toward the testing^16^, and 3) each provider completed a return of results survey for every one of their enrolled patients to query whether they believed testing affected their clinical management.

### Variables

Variables including demographic characteristics, biochemical parameters, CKD status, co-morbidities, and medication lists were obtained from the EHR at baseline and at 1 year follow-up.

### Outcomes

Study outcomes included: 1) prevalence of uncontrolled hypertension associated with actionable DGIs at baseline, and 2) change in systolic blood pressure (SBP) and diastolic blood pressure (DBP) at 1 year follow-up. A DGI or “actionable” genotype was defined as the presence of at least one variant predicting reduced efficacy for an antihypertensive agent a subject was taking at the time of enrollment. Secondary outcomes included patient attitudes toward genetic testing and provider utilization as defined by the return of results survey.

### Statistical Analysis

Baseline data and survey responses were analyzed descriptively and provided as percentages for categorical variables, mean ± standard deviation (SD) for normally distributed variables, and median (25th and 75th percentile) for non-normally distributed variables. Comparisons of categorical variables were expressed as an odds ratio (OR) with 95% confidence interval (CI). For our primary outcome analyses, subjects were considered to have a relevant DGI if one or more of their genetic variants predicted reduced efficacy of their prescribed antihypertensives. DGIs were coded as a binary variable (present or absent). Evaluation of the relationship between DGIs and hypertension control was performed by χ^2^ test and adjusted for significant covariates using logistic regression. Sub-group analyses were performed for each individual drug-gene(s) pair. Variants predicting increased efficacy of antihypertensives were assessed in sub-group analyses. Change in blood pressure within each individual at 1 year was assessed by Paired Student’s t-test.

## RESULTS

### Participants

A total of 472 adult subjects were recruited and consented from outpatient nephrology clinics within the Indiana University Health system, the Eskenazi Health safety-net system, and associated outlying suburban clinics (Figure 1). Thirty-seven subjects withdrew from the study, most frequently due to personal reasons, an inability to follow-up during the coronavirus pandemic, or a genotyping failure for which they would not provide a repeat sample. The remaining 435 subjects were genotyped and completed baseline surveys. The characteristics of the overall population are given in Table 2. There were approximately equal numbers of female and male subjects and the average age was 58.1 ± 14.9 years old. The average body mass index (BMI) of 33.7 ± 8.4 kg/m^2^. The majority had baseline CKD stage 3 and 92.2% had an ICD10 diagnosis of hypertension (N = 401); however, only 382 subjects (87.8%) were treated with antihypertensive agents. The average number of antihypertensive agents prescribed was 2.7 ± 1.6 per subject. Angiotensin converting enzyme inhibitors (ACEIs) and ARBs (57.6%) or beta blockers (55.6%) were commonly prescribed. Common comorbidities included diabetes, heart disease, and sleep apnea. The overall mean SBP was 139.9 ± 22.1 mmHg and the mean DBP was 80.7 ± 12.0 mmHg. Of the 382 subjects on antihypertensives, 335 subjects completed 1 year follow-up.

**Table 2:** Baseline Characteristics of Subjects included in Overall Genotype-Analysis

| Characteristic | Entire Cohort<br>(N = 435) (%) | uHTN<br>(N=189) (%) | cHTN<br>(N=193) (%) | P value<br>uHTN v cHTN |
| --- | --- | --- | --- | --- |
| Female Sex | 220 (50.6) | 90 (47.6) | 97 (50.3) | 0.71 |
| Age (yr) | 58.08 ± 14.19 | 59.48 ± 13.92 | 57.87 ± 13.42 | 0.65 |
| BMI (kg/m<sup>2</sup>)<sup>a</sup> | 33.7 ± 8.4 | 33.75 ± 8.85 | 34.11 ± 7.87 | 0.62 |
| Race (%) |  |  |  | 0.004 |
| White | 253 (58.2) | 92 (48.7) | 125 (64.8) |  |
| Black | 175 (40.2) | 93 (49.2) | 65 (33.7) |  |
| Asian | 6 (1.4) | 2 (1.1) | 4 (2.1) |  |
| Native American | 1 (0.2) | 1 (0.5) | 0 |  |
| Ethnicity |  |  |  |  |
| Hispanic | 6 (1.4) | 4 (2.1) | 2 (1.0) | 0.66 |
| *Baseline Blood Pressure (mm Hg)* |  |  |  |  |
| Average Systolic | 139.85 ± 22.06 | 157.96 ± 17.56 | 123.26 ± 9.64 | < 0.001 |
| Average Diastolic | 80.72 ± 11.98 | 86.86 ± 12.57 | 75.40 ± 8.66 | < 0.001 |
| *Chronic Kidney Disease Stage (%)* |  |  |  | 0.038 |
| I (eGFR >90 mL/min/1.73 m<sup>2</sup>) | 34 (7.82) | 16 (8.47) | 10 (5.18) |  |
| II (eGFR 60-89 mL/min/1.73 m<sup>2</sup>) | 40 (9.20) | 18 (9.52) | 18 (9.33) |  |
| III (eGFR 30-59 mL/min/1.73 m<sup>2</sup>) | 245 (56.32) | 94 (49.74) | 125 (64.77) |  |
| IV (eGFR 29 to 15 mL/min/1.73 m<sup>2</sup>) | 67 (15.40) | 39 (20.63) | 25 (12.95) |  |
| V (eGFR <15 mL/min/1.73 m<sup>2</sup>) | 26 (5.98) | 15 (7.94) | 9 (4.66) |  |
| No Kidney Disease | 23 (5.29) | 7 (3.70) | 6 (3.11) |  |
| *Coexisting Medical Conditions (%)* |  |  |  |  |
| Hypertension | 401 (92.18) | 189 (100) | 193 (100) | >0.99 |
| Diabetes | 198 (45.51) | 93 (49.21) | 83 (43.01) | 0.22 |
| Coronary Artery Disease/Prior MI | 72 (16.55) | 33 (17.46) | 35 (18.13) | 0.86 |
| Congestive Heart Failure | 71 (16.32) | 35 (18.52) | 29 (14.95) | 0.36 |
| Peripheral Vascular Disease | 22 (5.05) | 12 (6.35) | 7 (3.63) | 0.22 |
| Obstructive Sleep Apnea | 119 (27.36) | 51 (26.98) | 57 (29.53) | 0.58 |
| Cancer | 80 (18.39) | 31 (16.40) | 39 (20.21) | 0.34 |
| Non-steroidal anti-inflammatory use | 65 (14.94) | 31 (16.40) | 24 (12.44) | 0.27 |
| Current Smoking Status | 68 (15.63) | 33 (17.46) | 27 (13.99) | 0.35 |

<sup>a</sup>BMI is body mass index and was missing for 7 individuals. uHTN = Uncontrolled hypertension, blood pressure ≥ 140/90, cHTN = Controlled hypertension <140/90. Yr – year, eGFR – estimated glomerular filtration rate, MI – myocardial infarction

**Figure 1:**
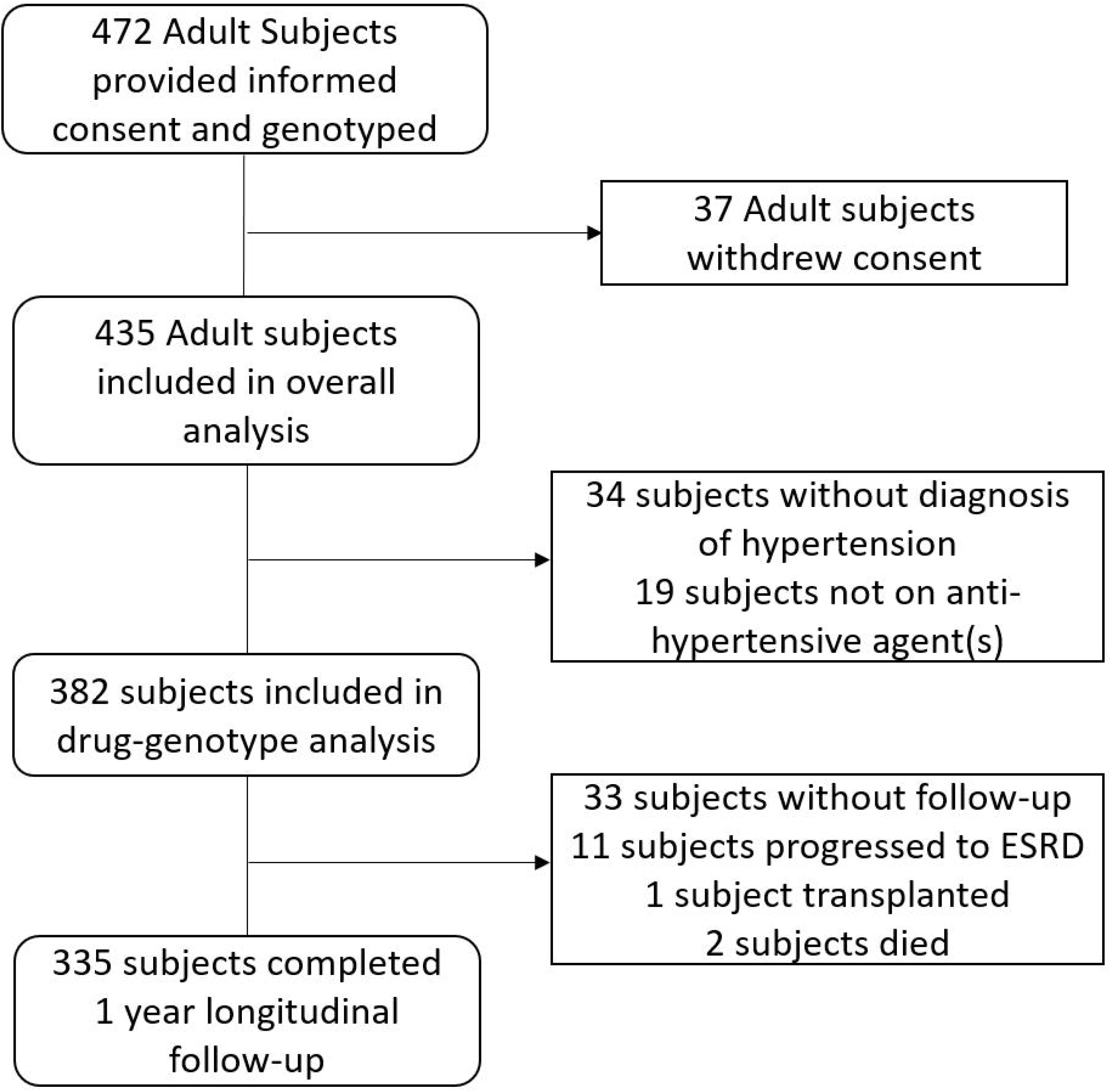
Enrollment and inclusion in the analyses. Survey data was available in 435 adult participants who received the pharmacogenomic genotyping panel. There were 382 subjects who had a hypertension diagnosis and were prescribed 1 or more antihypertensives. These 382 were included in the cross-sectional analysis. In the longitudinal analysis, there were 335 subjects included who completed a 1 year follow-up with subsequent blood pressure assessment.

### Association of drug-gene interactions and blood pressure control

Of the 382 individuals prescribed antihypertensive agents at baseline, 189 had uncontrolled hypertension (uHTN), defined as a SBP ≥ 140 mm Hg or DBP ≥ 90 (Table 2). The mean SBP in subjects with uHTN was 158.0 ± 17.6 mmHg. The mean SBP in individuals with controlled hypertension (cHTN, N = 193) was significantly lower at 123.3 ± 9.7 (P < 0.001). Similarly, the average DBP in uHTN subjects was higher (P < 0.001). Those with cHTN and uHTN were similar in age, sex distribution, BMI, and frequency of comorbidities. CKD was more prevalent in the uHTN group (P = 0.038). The distribution of race was significantly different between groups as 64.8% of cHTN subjects were white while only 48.7% of uHTN subjects were white (P = 0.004).

In the 382 subjects at baseline, relevant DGIs predicted to reduce efficacy of a currently prescribed antihypertensive agent were assessed. The genotype distribution is summarized in Supplemental Table S2. The majority of participants with uHTN had an actionable genotype (58.2%). Subjects with relevant DGIs were more likely to have uncontrolled hypertension (P = 0.0008). Subjects with an actionable genotype had 2-fold (95% CI 1.3-3.0) higher odds of uHTN as compared to those without an actionable genotype (Table 3). When adjusted for race and presence of CKD stage 3 or greater, subjects with a relevant DGI had 1.88-fold (95% CI 1.2-2.8) increased odds of uHTN. In summary, individuals who had uHTN were more likely to have a genetic variant that predicted reduced efficacy of an anti-hypertensive agent that they were prescribed at baseline.

**Table 3:** Association of antihypertensive drug-gene interactions with blood pressure

| Group | uHTN<br>N = 189 (%) | cHTN<br>N = 193 (%) | Odds Ratio (95% CI) | P value |
| --- | --- | --- | --- | --- |
| Unadjusted $\chi^2$ analysis | | | | |
| Relevant DGI<sup>A</sup> | 110 (58.2) | 79 (40.9) | 2.01 (1.3-3.0) | 0.0008 |
| No relevant DGI | 79 (41.8) | 114 (59.1) |  |  |
| Logistic Regression analysis |  |  |  |  |
| Relevant DGI |  |  | 1.88 (1.2-2.8) | 0.0005 |
| Race (non-white) |  |  | 1.45 (1.04-2.0) |  |
| Chronic Kidney Disease (eGFR < 60 ml/min) |  |  | 0.71 (0.4-1.2) |  |

<sup>A</sup>Relevant DGI (drug gene interaction) refers to the presence of a reduced efficacy variant for an antihypertensive agent a subject was taking on enrollment. uHTN – uncontrolled hypertension, cHTN – controlled hypertension, CI – confidence interval. Unadjusted analysis performed with a $\chi^2$ analysis. Logistic regression analysis was adjusted for race and stage of CKD.

### Individual drug-gene analyses

As exploratory analyses, we examined the association between relevant DGIs and baseline uHTN for each individual drug-gene interaction (Supplemental Table 3). Significant associations between uHTN and DGIs were found for participants prescribed losartan, metoprolol, and carvedilol. Variants in *CYP2C9* that predicted reduced efficacy of losartan were associated with uHTN in participants taking the drug (OR 5.2, 95% CI 1.9 to 14.7). Intermediate or poor CYP2D6 metabolizers have higher circulating concentrations of metoprolol or carvedilol. These individuals were less likely to have uncontrolled hypertension than normal metabolizers taking either agent (OR 0.55, 95% CI 0.3-0.95). No other significant DGIs were identified in this relatively small sample size.

### Longitudinal blood pressure control

Overall, the 335 subjects who completed a 1 year follow-up in the Nephrology clinic had a significant decrease in blood pressures, both systolic (−4.0 (95% CI 1.6,6.5) mmHg) and diastolic (−3.3 (95% CI 2.0,4.6) mmHg). Amongst the 160 individuals with uHTN, 71 were “controlled” by 1 year follow-up with a SBP < 140 and a DBP < 90 mmHg. Table 4 shows the within group comparisons of baseline and 1 year follow-up blood pressures in the overall cohort, in those with a DGI, in those with uHTN at baseline and with uHTN and a concurrent actionable genotype. All comparisons were significant (p<0.001). The magnitude of reductions in both SBP at 14.8 (95%CI 10.3, 19.3) mmHg and DBP at 8.4 (95%CI 5.9,10.9) mmHg were largest in the 90 individuals with uHTN and an actionable genotype. Taken together, the inference is that the detection of an actionable genotype provided an opportunity for control of blood pressure over one year in those uncontrolled at baseline.

**Table 4:** Longitudinal Blood Pressure assessment

| Group | SBP BL | SBP 1 yr | Change (95% CI) | P | DBP BL | DBP 1 yr | Change (95% CI) | P |
| --- | --- | --- | --- | --- | --- | --- | --- | --- |
| All subjects<br>N = 335 | 140.2 ± 22.5 | 136.2 ± 19.8 | -4.0 (1.6, 6.5) | 0.002 | 80.8 ± 12.2 | 77.6 ± 11.3 | -3.3 (2.0, 4.6) | 8.6 × 10<sup>-7</sup> |
| DGI subjects<sup>A</sup><br>N = 160 | 143.2 ± 22.6 | 138.3 ± 18.9 | -4.8 (1.3, 8.3) | 0.008 | 81.5 ± 12.8 | 77.1 ± 11.1 | -4.4 (2.5, 6.3) | 1.0 × 10<sup>-5</sup> |
| uHTN<br>subjects<br>N = 163 | 157.9 ± 17.9 | 143.3 ± 20.9 | -14.1 (10.4, 17.8) | 3.0 × 10<sup>-13</sup> | 86.7 ± 12.5 | 79.4 ± 12.3 | -7.1 (5.2, 9.1) | 9.6 × 10<sup>-9</sup> |
| uHTN + DGI<sup>A</sup><br>subjects<br>N = 90 | 158.8 ± 16.9 | 144.0 ± 19.5 | -14.8 (10.3, 19.3) | 1.3 × 10<sup>-9</sup> | 86.1 ± 13.7 | 77.7 ± 11.6 | -8.4 (5.9, 10.9) | 4.1 × 10<sup>-9</sup> |

<sup>A</sup>Relevant DGI (drug gene interaction) refers to the presence of a reduced efficacy variant for an antihypertensive agent a subject was taking on enrollment. uHTN – uncontrolled hypertension, BL – baseline, SBP – systolic blood pressure, DBP – diastolic blood pressure, yr - year

### Provider utilization

Action driven item surveys for each recruited subject were completed by their primary nephrology provider. The surveys queried the utility of genotype results in each subjects’ anti-hypertensive drug management. The response rate was 53.7% (180 of 335). Physicians reported that the genetic testing altered their diagnosis or management in 36% of cases. In 85% of survey responses, physicians stated they had or would discuss results with their patients.

### Patient reported attitudes to genotyping

Among the full cohort of 435 subjects with hypertension, proteinuria, or eGFR < 60 mL/min/1.73 m^2^, 425 subjects completed a baseline survey. Supplemental Figure 1 illustrates the response from selected questions regarding subjects’ attitude toward genetic testing. The surveys revealed that few subjects (5.4%) were familiar with the terms “pharmacogenomics, genetic testing or personalized medicine”. More than 96% reported that knowledge of their genetic code would prompt them to invest more to control their blood pressure as well as enable their providers to deliver enhanced antihypertensive care.

## DISCUSSION

Antihypertensive medication response is frequently unpredictable and varies among individuals. In CKD, blood pressure control at recommended targets is clearly important to prevent cardiovascular disease and end organ dysfunction. However, 24.9 to 33.4% of individuals with grade 3 or greater CKD have uncontrolled BP despite therapy with three or more medications^17^. Society guidelines often provide initial and secondary agent recommendations extrapolated from the general population^18, 19^, but may fail to provide specific direction for those with CKD and apparent treatment resistant (uncontrolled) hypertension.

In this study, relevant drug-gene interactions were found in 58.2 % of subjects. We identified a significant odds ratio of 1.8 between reduced efficacy drug-gene interactions and uncontrolled hypertension, even after adjusting for presence of CKD and race. When providers were armed with and trained on the genotype information, they reported adjusting the antihypertensive regimen in 36% of subjects. Study subjects were receiving ongoing care in nephrology clinics including frequent follow-up, ambulatory BP monitoring when required, secondary HTN work-up, and dietary counseling. The only addition to standard care was the provision of the genotype report in the electronic health record and notification to the subject’s physician. Medication changes, if any, were made by the primary nephrologists who indicated they discussed the genetic results with 85% of subjects. Patient engagement is a necessary component of hypertension management. Indeed, surveyed subjects agreed that understanding their pharmacogenomic report would facilitate greater action on their part to control BP. The adjustments made by the patients and providers resulted in a decrease of 4 mm Hg in SBP within individuals across the entire cohort. A remarkable reduction in SBP of 14.8 mm Hg was observed in those with baseline uHTN and an actionable genotype.

Our pharmacogenomics panel-based approach included a wide array of variants. A summary of the available evidence is beyond the scope of this discussion; however, antihypertensive variants were selected based on the strength of evidence, minor allele frequency, FDA label annotations, and guidelines. The FDA drug labels of hydralazine^20^, losartan^8^, and metoprolol^21^ each reference metabolic enzymes which affect concentrations of these drugs. Where major society guidelines were available, such as for metoprolol and carvedilol from the Dutch Pharmacogenomic Working Group (DPWG)^22^, they were used to guide recommendations. As additional guidelines from DPWG and the Clinical Pharmacogenomics Implementation Consortium (CPIC) are made available, these will need to be incorporated into future genotype-guided dosing recommendations.

We included 40 variants associated with efficacy in a breadth of agents to maximize actionability and impact for enrolled subjects. In some cases, pharmacokinetic and pharmacodynamic variants predicted opposite effects for the same drug. For example, the CYP2D6 poor metabolizer status predicts increased circulating concentrations of metoprolol, yet a subject with a reduced function variant of *ADRB1* would have the opposite predicted efficacy. There is insufficient evidence to reconcile these two effects; thus, all variants were reported separately and the prescribing clinician decided how they would use the information. Consideration was given to evidence in multiple populations. The Pharmacogenomics Evaluation of Antihypertensive Response (PEAR) group identified different predictors of thiazide efficacy in African Americans and Caucasians. As such, *YEATS4* was used as a predictor in individuals with African ancestry^23^ while a different three gene model was used in Caucasians^24^. In this study, the outcomes of utilization and blood pressure control incorporate a level of decision making on the part of the provider. The direction of effect for drug-gene prediction, the identification of race by the provider and subject, and the choice of medications to treat apparent treatment resistant hypertension are inherent aspects of the decision-making process for all providers.

A significant strength of our study is generalizability. Our study population was 57% white, 40% black and recruited from three diverse environments: a university hospital, a safety net health system, and several suburban clinics. The primary limitation of this study is that it was a prospective cohort, not a randomized controlled trial. Thus, observation bias in BP control (the Hawthorne effect) may contribute to the outcomes measured. This bias is inherent in all prospective cohort studies. An additional weakness, although also more generalizable, is that we did not mandate specific changes to the BP regimen. Multiple prior studies have^11^. We were underpowered to detect the significance of specific prescribing behavior. In practice, this may hold less relevance as CKD patients are on multiple antihypertensives, frequently with multiple actionable genotypes. Some providers would elect to change dose, select an alternate medication, or add a medication. We relied upon a binary variable of provider self-report of utility in each subject. A pharmacogenomic panel-based approach was utilized, and our sample size was insufficient to detect the effect of any single variant. Significant loss to follow-up (8.6%) did occur, in part due to the advent of the coronavirus pandemic which shifted follow-up to telehealth and impacted assessment of BP. Finally, we did not track medication compliance. These limitations are counterbalanced by our clinically translatable outcomes such as the association of drug-gene interactions with BP, longitudinal BP control, and provider utilization.

This study outlines the implementation of pharmacogenomic panel testing in an outpatient nephrology setting. In our cohort, we showed that nephrology providers will use information regarding drug-gene interactions to effect change in blood pressures in their patients. Currently, individual patient demographics such as obesity, gender, and race are important factors in the selection process for an adequate anti-hypertensive regimen^25^. This study illustrates that there is a convincing role for the addition of pharmacogenomic data to make choices in antihypertensive regimens. We expect that in the future, whole genome or exome sequencing will be integrated into the clinical setting. At that time, genotype information will be readily available to providers and will usher in an exciting era in the care of patients with hypertension and chronic kidney disease.

## Data Availability

There is no publicly available data source

## Author contributions

Conception, study design: MTE, RNM, ABC, TCS, SMM Analysis, interpretation of data: MTE, RNM, JM, SMM, RMF, JS. Conduct of study: VMP, MTE Drafting / revising the article: MTE, RNM, JM. Providing intellectual content: ADS, BWM, SJS. Final approval of the version to be published: All authors.

## Funding

Support for this work was provided by the NIH/NIDDK K08DK107864 (M.T.E.) and the Indiana University Grand Challenge Precision Health Initiative. SMM was funded by R01DK11087103 and P30AR072581-01. RNM was supported by K23DK102824 and R01AR077273.

**Collaborators:** “for the CKD-PGX investigators”

Allon N. Friedman MD^1^, Kimberly S. Collins PhD^1^, Nehal A. Sheth BA^1^, Katherine M. Spiech BS^1^, Asif A. Sharfuddin MBBS^1^, Nupur Gupta MBBS^1^, Ayman Hallab MD^1^, Simit Doshi MBBS MPH^1^, Matthew D. Dollins MD^1^, Emma M. Tillman PharmD PhD^1^, Elizabeth Rowe PhD^1^, Tyler Shugg PharmD PhD^1^, Chad A. Zarse MD^1^, Jonathan W. Bazeley MD MS^1^, Jay B. Wish MD^1^, David S. Hains MD^2^, Myda Khalid MBBS^2^, Tae-Hwi Schwantes-An PhD^3^, Elizabeth B. Medeiros MSFS^3^

^1^Department of Medicine, Indiana University School of Medicine, Indianapolis, Indiana, 46202, USA ^2^Department of Pediatrics, Indiana University School of Medicine, Indianapolis, Indiana, 46202, USA ^3^Department of Medical and Molecular Genetics, Indiana University School of Medicine, Indianapolis, Indiana, 46202, USA

## Figure Legends

**Supplemental Figure 1: Subject reported survey results**. Of the 435 subjects who underwent genotyping, 425 completed a baseline survey regarding their beliefs and attitudes toward genetic testing. All responses were given according to a Likert scale. A) Subjects were asked about their familiarity with pharmacogenomics. B) Subjects were asked whether genetic testing would increase their own efforts to control their blood pressure. C-D) Subjects were asked whether the genetic testing would help their providers select medications and control blood pressure.

