## Supplemental Document for "Pharmacogenomics of hypertension in chronic kidney disease: the CKD-PGX study"

Supplemental Document Table of Contents

Page 2 – Supplemental Figure 1

Page 3 – Supplemental Table 1

Page 4 – Supplemental Table 2

Page 5 – Supplemental Table 3

Page 6 – Supplemental Document (Subject Survey)

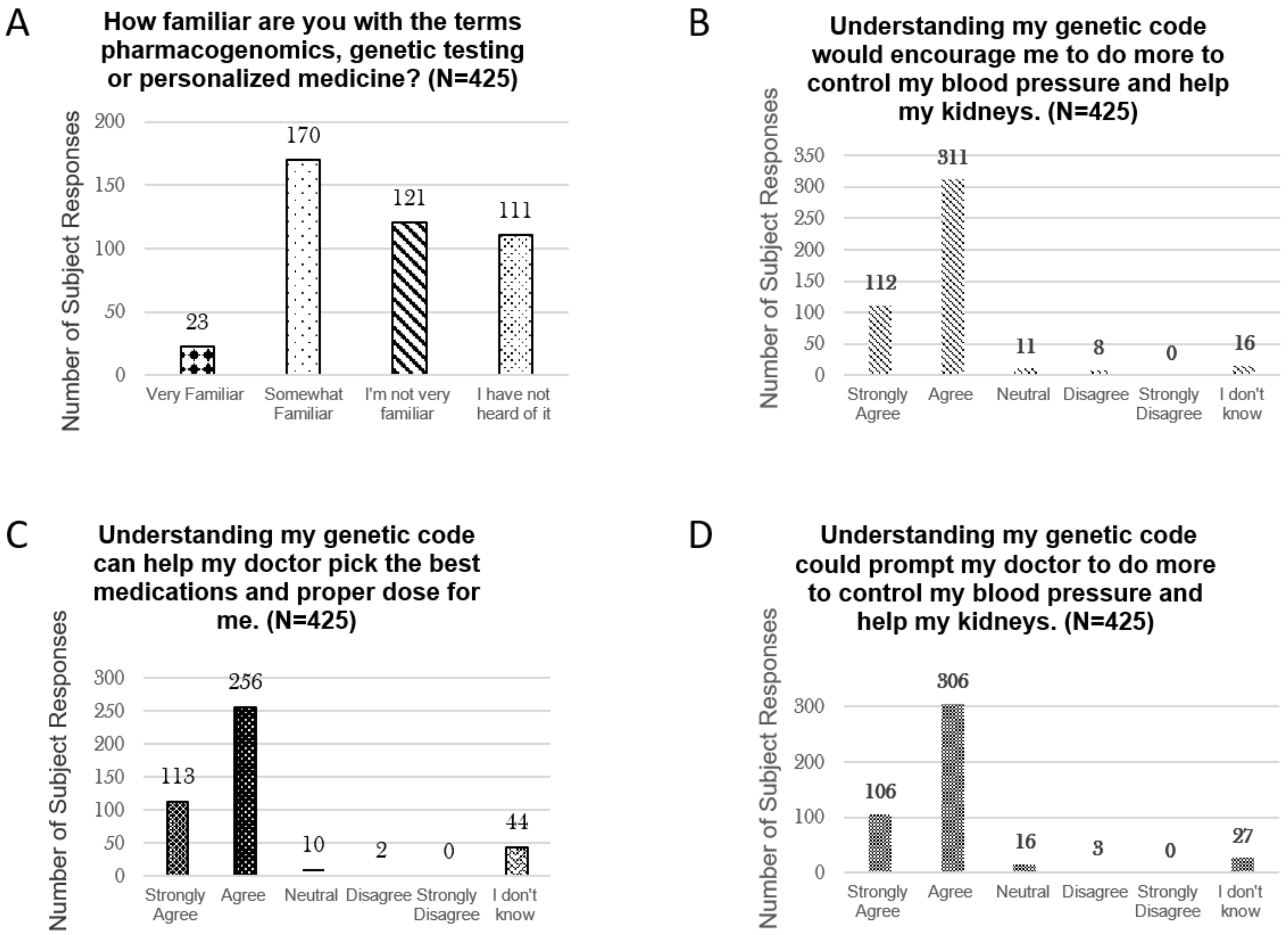


**Supplemental Figure 1: Subject reported survey results.** Of the 435 subjects who underwent genotyping, 425 completed a baseline survey regarding their beliefs and attitudes toward genetic testing. All responses were given according to a Likert scale. A) Subjects were asked about their familiarity with pharmacogenomics. B) Subjects were asked whether genetic testing would increase their own efforts to control their blood pressure. C-D) Subjects were asked whether the genetic testing would help their providers select medications and control blood pressure.

### Supplemental Table 1: Gene descriptions

| Gene | Variants Tested | Biological/Functional Significance | Predicted Phenotype |
|---|---|---|---|
| *ADRB1* | rs1801253 (c.1165G>C), rs1801252 (c.145A>G) | Encodes $\beta$1-adrenoreceptor, polymorphism is associated with greater agonist-promoted downregulation and altered glycosylation of the receptor. | Increased beta-blocker efficacy with increasing copies of the 49S-389R haplotype. |
| *CYP2C9* | CYP2C9*2 (c.430C>T), CYP2C9*3 (c.1075A>C), CYP2C9*5 (c.1080C>G)<br>CYP2C9*6 (c.818delA), CYP2C9*8 (c.449G>A), CYP2C9*11 (c.1003C>) | Encodes cytochrome P450 2C9 enzyme, which metabolizes losartan to its active metabolite E3179. E3179 is 10- to 40-fold more potent than its parent compound. | Intermediate or poor metabolizers have reduced efficacy of losartan. |
| *CYP2D6* | CYP2D6*2 (c.2850C>T and c.4180G>C), CYP2D6*3 (c.2549delA CYP2D6*4 (c.1846G>A and c.100C>T), CYP2D6*5 (gene deletion)<br>CYP2D6*6 (c.1707delT), CYP2D6*7 (c.2935A>C)<br>CYP2D6*9 (c.2615_2617delAAG), CYP2D6*10 (c.100C>T and c.4180G>C)<br>CYP2D6*17 (c.1023C>T), CYP2D6*29 (c.3183G>A), CYP2D6*41 (c.2988G>A), CYP2D6*1XN (duplication), CYP2D6*2XN (c.2850C>T, c.4180G>C, duplication) CYP2D6*4XN (c.1846G>A, c.100C>T, duplication) | Encodes cytochrome P450 2D6 enzyme, metabolizes metoprolol and carvedilol to its inactive metabolites. | Poor metabolizers have higher circulating concentrations and increased risk of bradycardia. Ultrarapid metabolizers have a risk of inefficacy. |
| *F7* | rs6046 (c.1238G>A; p.Arg413Gln) | Encodes vitamin K dependent clotting factor VII. F7 is the initiator of the extrinsic coagulation pathway. Blood pressure effect possibly related to endothelial homeostasis. | Decreased amlodipine efficacy with increasing copies of A allele in African Americans. |
| *GRK4* | rs2960306 (c.194G>T), rs1024323 (c.329C>G) | Encodes G-protein coupled receptor kinase 4 (GRK4). The presence of GRK4 is important for regulation of ADRB1 cell surface expression. | Decreased beta-blocker efficacy associated with increasing copies of the 65L-142V haplotype. |
| *NAT2* | NAT2*5 [rs1801280 (c.341T>C)], NAT2*6 [rs1799930 (c.590G>A)], NAT2*7 [rs1799931 (c.857G>A)], NAT2*14 [rs1801279 (c.191G>A)] | Encodes N-acetyletransferase 2, which is responsible for acetylation of hydralazine to its inactive metabolite. | Poor metabolizers have higher circulating concentrations of hydralazine and increased efficacy. |
| *NEDD4L* | rs4149601 (c.24G>A) | Encodes member of the Nedd4 family of HECT domain E3 ubiquitin ligases, regulating cell surface expression of the epithelial sodium channel, ENaC. Consists of 3 domains (C2, WW and Hect). C2 domain is a calcium-rich lipid-binding domain responsible for membrane targeting. WW and Hect domains target ENac for internalization into the cell and degradation by lysozomes respectively. | Presence of two G alleles results in increased ENaC activity and increased response to diuretics in Caucasians. |
| *NPHS1* | rs3814995 (g.35851310C>T) | Encodes nephrin, the principal structural protein of the glomerular podocytes. NPHS1 mutations result in congenital nephrotic syndrome. Angiotensin receptor blocks decrease renal nephrin expression. | One or more A alleles is associated with increased angiotensin receptor blocker efficacy. |
| *VASP* | rs10995 (c.*719G>A) | Encodes vasodilator-stimulated phosphoprotein (VASP). VASP is a substrate for cyclic AMP-and cyclic GMP-dependent protein kinases, regulating smooth muscle contraction. | Increased expression is associated with increased thiazide efficacy. |
| *YEATS4* | rs7297610 (g.69430244C>T) | Encodes GAS41, essential for RNA transcription and cell viability including chromatin-modification. Evidence for direct functional/biological significance lacking, possible role with regards to hypertension-associated cell proliferation at the level of the renal tubules. | One or more copies of the T allele is associated with decreased thiazide efficacy. |
| *EBF1/ FGF5/ SH2B3* | EBF1 rs4551053 (g.158411534G>A)<br>FGF5 rs1458038 (g.81164723C>T)<br>SH2B3 rs3184504 (c.784T>C) | This 3 gene model was uncovered in the PEAR studies and evidence for direct functional/biological significance is lacking. | Increasing copies of efficacy alleles are associated with increased thiazide efficacy. |

Supplemental Table 2: Genotype frequency

| Gene | Actionable Genotype | Overall Frequency (N = 382) |
| --- | --- | --- |
| *ADRB1* | Greater beta blocker response (2 copies of 49S-389R) | 42 (11.0) |
| | Standard beta blocker response (1 copy of 49S-389R) | 200 (52.4) |
| | Reduced beta blocker response (0 copies of 49S-389R) | 150 (39.3) |
| *CYP2D6* | Ultrarapid metabolizer | 12 (3.1) |
| | Normal or indeterminate metabolizer | 212 (55.5) |
| | Intermediate (Reduced) metabolizer | 141 (36.9) |
| | Poor metabolizer | 17 (4.5) |
| *CYP2C9* | Normal Metabolizer | 272 (71.2) |
| | Intermediate (Reduced) metabolizer (*1/*2) | 49 (12.8) |
| | Intermediate (Reduced) metabolizer (non-*1/*2) | 43 (11.2) |
| | Poor Metabolizer | 18 (4.7) |
| *F7* | Standard efficacy (G/G allele) | 305 (79.8) |
| | Reduced efficacy (G/A or A/A allele) | 77 (20.2) |
| *GRK4* | Greater beta blocker response (0 copies of 65L-142V) | 166 (43.5) |
| | Standard beta blocker response (1 copy of 65L-142V) | 174 (45.5) |
| | Reduced beta blocker response (2 copies of 65L-142V) | 42 (11.0) |
| *NAT2* | Normal or intermediate metabolizer | 180 (47.1) |
| | Poor metabolizer | 202 (52.9) |
| *NEDD4L* | Increased diuretic efficacy (G/G) | 171 (44.8) |
| | Standard diuretic efficacy (G/A) | 173 (45.3) |
| | Reduced diuretic efficacy (A/A) | 38 (9.9) |
| *NPHS1* | G/G (Standard ARB efficacy) | 243 (63.6) |
| | G/A or A/A (Increased ARB efficacy) | 139 (36.4) |
| *VASP* | Standard thiazide efficacy (A/A) | 236 (61.8) |
| | Increased efficacy allele (A/G or G/G) | 146 (28.2) |
| YEATS4 | Standard efficacy (C/C) | 266 (69.6) |
| | Reduced efficacy allele (C/T or T/T) | 116 (30.4) |
| *EBF1/FGF5/SH2B3* | Increased thiazide efficacy (3 or more efficacy alleles) | 92 (24.1) |
| | Standard thiazide efficacy (1 or 2 efficacy alleles) | 281 (73.6) |
| | Reduced thiazide efficacy (0 efficacy alleles) | 9 (2.4) |

Supplemental Table 3: Cross sectional associations of individual drug-gene interactions with baseline uncontrolled hypertension

| *Drug-Gene Interaction* | *Predicted direction of effect* | *uHTN* | *cHTN* | *Odds Ratio (CI)* | *P-value* | *N*[D] |
|---|---|---|---|---|---|---|
| ***F7*-amlodipine**[A] | Normal variant | 48 | 38 | 1.98 (0.8-5.0) | 0.15 | 114 |
| | Reduced efficacy | 20 | 8 | | | |
| ***NEDD4L*-diuretics**[A] | Normal / increased | 23 | 45 | 1.83 (0.9-3.7) | 0.09 | 130 |
| | Reduced efficacy | 30 | 32 | | | |
| ***ADRB1*-beta blockers** | Normal / increased | 68 | 74 | 1.47 (0.9-2.5) | 0.15 | 236 |
| | 2 Reduced efficacy alleles | 54 | 40 | | | |
| ***GRK4*-beta blockers** | Normal Variant | 107 | 102 | 1.19 (0.5-2.7) | 0.67 | 236 |
| | Reduced efficacy | 15 | 12 | | | |
| ***CYP2C9*-losartan**[B] | Normal | 39 | 88 | **5.2 (1.9-14.7)** | **0.002** | 108 |
| | Intermediate / Poor | 14 | 6 | | | |
| ***CYP2D6*-beta blockers**[C] | Normal | 79 | 55 | **0.55 (0.3-0.95)** | **0.03** | 222 |
| | Intermediate/ Poor | 39 | 49 | | | |
| ***NPHS1*-ARB** | Normal Variant | 35 | 33 | 0.77 (0.4-1.7) | 0.52 | 108 |
| | Increased efficacy allele | 18 | 22 | | | |
| ***VASP*-Thiazides**[A] | Normal Variant | 7 | 14 | 0.86 (0.3-2.8) | 0.80 | 51 |
| | Increased efficacy allele | 9 | 21 | | | |

[A]Denotes the analysis was performed in the relevant population. *F7* was tested in individuals with self-reported African ancetsry. *NEDD4L* and *VASP* were tested in individuals of self-reported European ancestry. [B]*CYP2C9* *1/*2 genotypes were included with normal metabolizers. [B]Ultrarapid and indeterminate metabolizers were excluded from the analysis. [D]Comparisons with sample size below 50 not shown.

**Supplemental Document 1:** Subject Survey

Study ID number: ______________ Date: _________________

Interviewer: ______________________

**Genetics Opinion Survey – Initial (Read to subject)**
By completing this survey you are agreeing to participate in a research study looking at your opinion on genetic testing. This information may be used to make education materials that can be helpful for you and other patients.

How familiar are you with the terms pharmacogenomics, genetic testing or personalized medicine?
- Very familiar
- Somewhat familiar
- Not very familiar
- I have not heard of it

Have you, or has anyone in your immediate family, ever been told that you carried a gene that predisposed you to certain diseases?
- Yes
- No
- I don't know

If a genetic test was available that could tell whether a family member was at higher risk to have their kidneys stop working in the future, would you want them to take the test?
- Yes
- No
- I don't know

Do you believe tests that use genes to predict diseases are mostly accurate and reliable?
- Yes
- No
- I don't know

<u>For each of the following statements, indicate whether you agree or disagree with them:</u>
Understanding my genetic code can help my doctor pick the best medications and proper dose for me.
- Strongly Agree
- Agree
- Neutral
- Disagree
- Strongly Disagree
- I don't know

Understanding my genetic code *would encourage me* to do more to control my blood pressure and protect my kidneys.

- Strongly Agree
- Agree
- Neutral
- Disagree
- Strongly Disagree
- I don't know

Understanding my genetic code could prompt my doctor to do more to control my blood pressure and help my kidneys.

- Strongly Agree
- Agree
- Neutral
- Disagree
- Strongly Disagree
- I don't know

How would you describe your health?

- Excellent
- Very Good
- Good
- Fair
- Poor

How would you describe your race and/or ethnicity?

- White
- African-American or Black
- Hispanic
- Asian
- Native American
- Other: ____________

How would you describe your highest level of education?

- Some High School (or less)
- High School Graduate or GED
- Some College (no degree)
- Bachelor's degree
- Graduate School (or higher)
